# Knowledge, attitudes and perceptions towards COVID-19 vaccinations: a cross-sectional community survey in Bangladesh

**DOI:** 10.1101/2021.02.16.21251802

**Authors:** Md. Saiful Islam, Abu Bakkar Siddique, Rejina Akter, Rafia Tasnim, Md. Safaet Hossain Sujan, Paul R Ward, Md. Tajuddin Sikder

## Abstract

**Background:** Several vaccines have been approved against coronavirus disease (COVID-19) and distributed globally in different regions. However, general community knowledge, attitudes and perceptions towards COVID-19 vaccinations are poorly understood. Thus, the study aimed to investigate community knowledge, attitudes and perceptions towards COVID-19 vaccinations in Bangladesh.

**Methods:** An exploratory and anonymous population-based e-survey was conducted among 1658 general individuals (55.6% male; mean age=23.17±6.05 years; age range=18-65 years). The survey was conducted using a semi-structured and self-reported questionnaire containing informed consent along with four sections (i.e., socio-demographics, knowledge, attitudes, and perceptions). Multiple linear regression was performed to determine the variables predicting knowledge, and attitudes towards COVID-19 vaccinations.

**Results:** The mean scores of knowledge and attitudes were 2.83±1.48 (out of 5) and 9.34±2.39 (out of 12) respectively. About a quarter of participants thought that the COVID-19 vaccination available in Bangladesh is safe, only 60% will have the vaccination and about two-thirds will recommend it to family and friends. In the multiple regression model, higher SES, having university/ higher levels of education, holding nuclear families and having previous history of essential vaccines uptake were associated with knowledge; whilst attitudes were significantly associated with being female and having previous history of essential vaccines uptake. Just over half of the participants thought that everyone should be vaccinated and 61% responded that health workers should be vaccinated first on priority basis. 95% vaccine should be administered free of charge in Bangladesh and almost 90% believed that the COVID-19 vaccine used in Bangladesh may have side effects.

**Conclusions:** The findings reflect inadequate knowledge but more positive attitudes towards COVID-19 vaccine among the general population in Bangladesh. In order to improve knowledge, immediate health education programs need to be initiated before mass vaccination schedule.

## Introduction

Coronavirus disease (COVID-19) is a deadly disease which continues to affect many countries in the world. This is caused by the new coronavirus strain SARS-CoV-2 which has become a serious public health concern worldwide[1]. The World Health Organization (WHO) declared the COVID-19 outbreak as a pandemic on 11 March 2020 [2]. At the time of writing (6 February 2021), this pandemic has affected 223 countries, with over 104.37 million confirmed cases and 22.71 million deaths recorded globally [3]. The incidence is higher in the Americas (46313540 cases and 1072244 deaths) and Europe (35003091 cases and 767235 deaths) than in South East Asia (12982540 cases and 199668 deaths), Africa (2616892 cases and 64473 deaths) and the Western Pacific (1466248 cases and 25526 deaths) [3]. The first COVID-19 case in Bangladesh was reported on 8 March 2020 [4,5]. Since then, the numbers of new cases have been rising rapidly in the country. As of 6 February 2021, the country has recorded 537465 positive cases of COVID-19 and 8182 deaths domestically [6].

Vaccines are the most important public health measure and most effective strategy to protect the population from COVID-19, since SARS-CoV-2 is highly contagious virus and affects populations widely and globally. The competition for COVID-19 vaccine invention and development against the spread and catastrophic effects of the disease is ongoing [7,8], and new, more effective vaccines are likely to be developed as we move through the pandemic. With the distribution of vaccines underway, it is very important to examine community acceptance of COVID-19 vaccinations [9]. Bangladeshi authorities agreed to use the Covidshield vaccine from India on 27 January 2021 and Runu (A Nurse) became the first COVID-19 vaccine recipient [10]. But there is a great controversy about COVID-19 vaccinations among the general population of Bangladesh. A proportion of the Bangladeshi population are hesitating to take the vaccine from India lest they should be infected [11]. A global survey of potential COVID-19 vaccine acceptance shows that 48% of their study population were confused about the COVID-19 vaccinations and remained unsure about whether they would have the vaccination [12]. Similarly, a Chinese study found that only just over half of their participants (54.%) said that they intended to have the vaccination [13]. These relatively low proportions of people willing to have the vaccine are potentially worrying, since although the most effective measure of controlling the spread of the virus is to protect oneself from being exposed to COVID-19, it is also necessary to vaccinate the vulnerable group of people as soon as possible [14].

In order implement the most effective vaccination strategy in Bangladesh, we need to know the knowledge, attitudes and perceptions of Bangladeshi people about COVID-19 vaccinations. In such a scenario, people’s knowledge, attitudes and perceptions towards COVID-19 are crucial for Government and policy makers to address all barriers to vaccine distribution. To date, there has been no prior study among the general population of Bangladesh investigating their knowledge, attitudes and perceptions towards COVID-19 vaccine.

## Methods

### Participants and procedure

An exploratory and anonymous population-based e-survey was conducted among individuals aged above 18 years. The survey was conducted from December 2020 to February 2021, coinciding with the implementation of COVID-19 vaccinations programs in other parts of the world. Thus, rather then being a hypothetical study of knowledge, attitudes and perceptions, our study aimed to capture real-world evidence to inform policy makers and practitioners in Bangladesh in terms of how best to implement our vaccination program. A semi-structured questionnaire was designed and incorporated into the Google survey tool (Google Forms) and a shareable link was generated and disseminated publicly on various social media outlets (e.g., Facebook, WhatsApp, etc.). Likewise, the link was also shared personally to the contact lists of the investigators and research assistants. For data collection, the investigators used online approaches (as opposed to face-to-face data collection) because of maintaining spatial distancing and proper precaution during the pandemic. Initially, 1738 respondents provided informed consent via e-survey. Of these, 1658 respondents (55.6% males; mean age = 23.17 ± 6.05; age range = 18-65 years) completed the entire survey. The inclusion criteria of participants were i) being a Bangladeshi resident, ii) being adults (≥18 years old), iii) having good internet access, and iv) having voluntary participation. The exclusion criteria included being under 18 years old and incomplete surveys. The study was conducted following the Checklist for Reporting Results of Internet ESurveys (CHERRIES) guidelines [15].

### Sampling procedure

The sample size was calculated using the following equation (1):

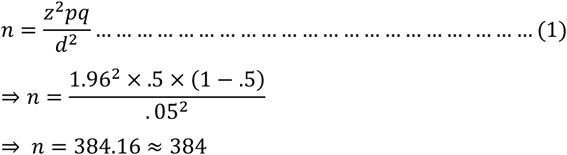

Here,

*n* = number of samples

*z* = 1.96 (95% confidence level)

*p* = prevalence estimate (50% or .5) (as no study found)

*q* = (1-*p*)

*d* = precision limit or proportion of sampling error (.05)

As there is no prior similar study focusing on KAP regarding the COVID-19 vaccine in Bangladesh, we made the best assumption (p) for the present study would be 50%. Assuming a 10% non-response rate, a sample size of 423.5 ≈ 424 participants was estimated. Our sample size exceeded this estimate.

### Measures

A semi-structured and self-reported questionnaire containing informed consent along with four sections (i.e., socio-demographics, knowledge, attitudes, and perceptions) was utilized during data collection (see questionnaire in **Supplementary file**).

#### Socio-demographic information

Some questions related to socio-demographics were asked during the survey including age, sex (male/female), marital status (married/unmarried), educational status (college/ below, university or higher), family type (nuclear [two parents and their children]/joint [family unit with more than two parents, extended family]; [16]), monthly family income (later categorized: lower socioeconomic status (SES) <15,000 Bangladeshi Taka [BDT], middle SES 15,000-30,000 BDT and upper SES >30,000 BDT) [17,18], and current residence (rural/urban). In addition, another “yes/no” question was asked about their previous history of taking all the recommended vaccines (i.e., *Have you received all the necessary vaccines in your lifetime?*).

#### Knowledge, attitudes, and perceptions

To assess the level of knowledge, attitudes, and perceptions of the respondents, a total of 19 items structured questions (including 6-items for knowledge, 6-items for attitudes and 6-items for perceptions). All questions were based on validated questions in previous literature [19–22].

The knowledge section comprised 5-items with three possible responses (i.e., *“Yes”, “No”* and *“Don’t know”*) (e.g., *Does vaccination increase allergic reactions?*) (see details in Table 2). The ‘*yes*’ response was coded as 1, while the ‘*No/ Don’t know*’ responses were conducted as 0. The total score was obtained by summating the raw scores of five items and ranged from 0-5, with the higher score indicating the greater level of knowledge towards COVID-19 vaccinations. The Cronbach alpha of knowledge items was 0.68. In addition, an additional question was asked about the source of their knowledge about COVID-19 vaccines (i.e., *How you came to know about COVID-19 vaccines first?*) with some possible answers (e.g., mass media [radio/television], newspaper, internet, social media [Facebook, Twitter], family and relatives, friends and neighbors). The answers to this question are useful for policy makers in terms of disseminating COVID-19 vaccination information in the future in Bangladesh.

**Table 1:** General characteristics of participants (N = 1658)

| **Variables** | **n** | **(%)** |
| --- | --- | --- |
| **Age** |  |  |
| Young | 1442 | (87.0) |
| Adult | 216 | (13.0) |
| **Sex** |  |  |
| Male | 922 | (55.6) |
| Female | 736 | (44.4) |
| **Marital status** |  |  |
| Unmarried | 1417 | (85.5) |
| Married | 241 | (14.5) |
| **Education** |  |  |
| College/ below | 289 | (17.4) |
| University/ higher | 1369 | (82.6) |
| **Family type** |  |  |
| Nuclear | 1318 | (79.5) |
| Joint | 340 | (20.5) |
| **Monthly family income** |  |  |
| Lower SES | 421 | (25.4) |
| Middle SES | 568 | (34.3) |
| Upper SES | 669 | (40.3) |
| **Residence** |  |  |
| Rural | 570 | (34.4) |
| Urban | 1088 | (65.6) |
| **Have you received all the necessary vaccines in your lifetime?** |  |  |
| Yes | 1225 | (73.9) |
| No | 433 | (26.1) |

**Table 2:** Distribution of each knowledge item and sex difference

| Variables | *Total* | | *Male* | | *Female* | | $\chi^2$ | df | *p*-value |
| --- | --- | --- | --- | --- | --- | --- | --- | --- | --- |
|  | n | (%) | n | (%) | n | (%) |  |  |  |
| **Do you know about the COVID-19 vaccine?** |  |  |  |  |  |  |  |  |  |
| Yes | 1492 | (90.0) | 830 | (90.0) | 662 | (89.9) | 1.97 | 2 | .373 |
| No | 93 | (5.6) | 56 | (6.1) | 37 | (5.0) |  |  |  |
| Don't know | 73 | (4.4) | 36 | (3.9) | 37 | (5.0) |  |  |  |
| **Do you know about the effectiveness of COVID-19 vaccine?** |  |  |  |  |  |  |  |  |  |
| Yes | 901 | (54.3) | 513 | (55.6) | 388 | (52.7) | 2.46 | 2 | .292 |
| No | 324 | (19.5) | 182 | (19.7) | 142 | (19.3) |  |  |  |
| Don't know | 433 | (26.1) | 227 | (24.6) | 206 | (28.0) |  |  |  |
| **Is it dangerous to use overdose vaccines?** |  |  |  |  |  |  |  |  |  |
| Yes | 1048 | (63.2) | 570 | (61.8) | 478 | (64.9) | 4.18 | 2 | .124 |
| No | 45 | (2.7) | 31 | (3.4) | 14 | (1.9) |  |  |  |
| Don't know | 565 | (34.1) | 321 | (34.8) | 244 | (33.2) |  |  |  |
| **Does vaccination increase allergic reactions?** |  |  |  |  |  |  |  |  |  |
| Yes | 656 | (39.6) | 384 | (41.6) | 272 | (37.0) | 5.88 | 2 | .053 |
| No | 78 | (4.7) | 48 | (5.2) | 30 | (4.1) |  |  |  |
| Don't know | 924 | (55.7) | 490 | (53.1) | 434 | (59.0) |  |  |  |
| **Does vaccination increase autoimmune diseases?** |  |  |  |  |  |  |  |  |  |
| Yes | 588 | (35.5) | 361 | (39.2) | 227 | (30.8) | 14.15 | 2 | .001 |
| No | 95 | (5.7) | 56 | (6.1) | 39 | (5.3) |  |  |  |
| Don't know | 975 | (58.8) | 505 | (54.8) | 470 | (63.9) |  |  |  |

The attitude section consisted of 6-items (e.g. *The newly discovered COVID-19 vaccine is safe; I will take the COVID-19 vaccine without any hesitation, if it is available in Bangladesh*.), and the response of each item was indicated on a three-point Likert scale (i.e., *0 = Disagree, 1 = Undecided*, and *2 = Agree*) (see details in Table 3). The total score was calculated by summating the raw scores of the six items ranging from 0 to 12, with an overall greater score indicating more positive attitudes towards COVID-19 vaccine. The Cronbach alpha of attitudes items was 0.77.

**Table 3:** Distribution of each attitudes item and sex difference

| Variables | Total | | Male | | Female | | $\chi^2$ | df | p-value |
| --- | --- | --- | --- | --- | --- | --- | --- | --- | --- |
|  | n | (%) | n | (%) | n | (%) |  |  |  |
| The newly discovered COVID-19 vaccine is safe. |  |  |  |  |  |  |  |  |  |
| Disagree | 78 | (4.7) | 50 | (5.4) | 28 | (3.8) | 14.60 | 2 | .001 |
| Undecided | 1157 | (69.8) | 608 | (65.9) | 549 | (74.6) |  |  |  |
| Agree | 423 | (25.5) | 264 | (28.6) | 159 | (21.6) |  |  |  |
| The COVID-19 vaccine is essential for us. |  |  |  |  |  |  |  |  |  |
| Disagree | 70 | (4.2) | 47 | (5.1) | 23 | (3.1) | 4.17 | 2 | .124 |
| Undecided | 363 | (21.9) | 196 | (21.3) | 167 | (22.7) |  |  |  |
| Agree | 1225 | (73.9) | 679 | (73.6) | 546 | (74.2) |  |  |  |
| I will take the COVID-19 vaccine without any hesitation, if it is available in Bangladesh. |  |  |  |  |  |  |  |  |  |
| Disagree | 168 | (10.1) | 113 | (12.3) | 55 | (7.5) | 11.43 | 2 | .003 |
| Undecided | 519 | (31.3) | 272 | (29.5) | 247 | (33.6) |  |  |  |
| Agree | 971 | (58.6) | 537 | (58.2) | 434 | (59.0) |  |  |  |
| I will also encourage my family/friends/relatives to get vaccinated. |  |  |  |  |  |  |  |  |  |
| Disagree | 112 | (6.8) | 80 | (8.7) | 32 | (4.3) | 12.25 | 2 | .002 |
| Undecided | 460 | (27.7) | 253 | (27.4) | 207 | (28.1) |  |  |  |
| Agree | 1086 | (65.5) | 589 | (63.9) | 497 | (67.5) |  |  |  |
| It is not possible to reduce the incidence of COVID-19 without vaccination. |  |  |  |  |  |  |  |  |  |
| Disagree | 225 | (13.6) | 154 | (16.7) | 71 | (9.6) | 17.86 | 2 | .000 |
| Undecided | 381 | (23.0) | 210 | (22.8) | 171 | (23.2) |  |  |  |
| Agree | 1052 | (63.4) | 558 | (60.5) | 494 | (67.1) |  |  |  |
| The COVID-19 vaccine should be distributed fairly to all of us. |  |  |  |  |  |  |  |  |  |
| Disagree | 31 | (1.9) | 21 | (2.3) | 10 | (1.4) | 2.00 | 2 | .369 |
| Undecided | 150 | (9.0) | 85 | (9.2) | 65 | (8.8) |  |  |  |
| Agree | 1477 | (89.1) | 816 | (88.5) | 661 | (89.8) |  |  |  |

The perceptions section included 6-items regarding participant’s perceptions towards the COVID-19 vaccine, including 4 items as “yes/no” questions (e.g., *Do you think the vaccine should be administered free of charge in Bangladesh?*) and additional 2 items related to the application of COVID-19 vaccine (e.g., *Which population groups should be prioritized for vaccinations?*) (see details in Table 4).

**Table 4:** Distribution of each perceptions item and sex difference

| Variables | *Total* | | *Male* | | *Female* | | $\chi^2$ | df | *p*-value |
| --- | --- | --- | --- | --- | --- | --- | --- | --- | --- |
|  | n | (%) | n | (%) | n | (%) |  |  |  |
| **Do you think the newly discovered COVID-19 vaccine may have side effects?** |  |  |  |  |  |  |  |  |  |
| Yes | 1477 | (89.1) | 822 | (89.2) | 655 | (89.0) | .011 | 1 | .918 |
| No | 181 | (10.9) | 100 | (10.8) | 81 | (11.0) |  |  |  |
| **Do you think that if everyone in the society maintains the preventive measures, the COVID-19 pandemic can be eradicated without vaccination?** |  |  |  |  |  |  |  |  |  |
| Yes | 930 | (56.1) | 527 | (57.2) | 403 | (54.8) | .960 | 1 | .327 |
| No | 728 | (43.9) | 395 | (42.8) | 333 | (45.2) |  |  |  |
| **Who should have been vaccinated, do you think?** |  |  |  |  |  |  |  |  |  |
| Those who have not yet been infected | 398 | (24.0) | 241 | (26.1) | 157 | (21.3) | 13.29 | 3 | .004 |
| People infected with COVID-19 | 374 | (22.6) | 224 | (24.3) | 150 | (20.4) |  |  |  |
| Newly recovered from COVID-19 | 25 | (1.5) | 15 | (1.6) | 10 | (1.4) |  |  |  |
| Everyone | 861 | (51.9) | 442 | (47.9) | 419 | (56.9) |  |  |  |
| **Who's supposed to be vaccinated first, you think?** |  |  |  |  |  |  |  |  |  |
| General public | 452 | (27.3) | 240 | (26.0) | 212 | (28.8) | 12.12* | 5 | .029 |
| Health worker | 1010 | (60.9) | 555 | (60.2) | 455 | (61.8) |  |  |  |
| Public/ private employee | 46 | (2.8) | 33 | (3.6) | 13 | (1.8) |  |  |  |
| Teacher/ student | 98 | (5.9) | 59 | (6.4) | 39 | (5.3) |  |  |  |
| Garment worker | 47 | (2.8) | 30 | (3.3) | 17 | (2.3) |  |  |  |
| Businessman | 5 | (.3) | 5 | (.5) | 0 | (.0) |  |  |  |
| **Do you think the vaccine should be administered free of charge in Bangladesh?** |  |  |  |  |  |  |  |  |  |
| Yes | 1577 | (95.1) | 868 | (94.1) | 709 | (96.3) | 4.22 | 1 | .040 |
| No | 81 | (4.9) | 54 | (5.9) | 27 | (3.7) |  |  |  |
| **Would you afford the vaccine at your own expense if it was not provided free by the government, do you think?** |  |  |  |  |  |  |  |  |  |
| Yes | 1084 | (65.4) | 593 | (64.3) | 491 | (66.7) | 1.04 | 1 | .308 |
| No | 574 | (34.6) | 329 | (35.7) | 245 | (33.3) |  |  |  |

\*Fisher's exact test

### Statistical analysis

The data analysis was performed using Microsoft Excel 2019 and SPSS version 25.0 (Chicago, IL, USA). Microsoft Excel was used for data cleaning, editing, sorting, and coding. The excel file was then imported into SPSS software. Descriptive statistics (i.e., frequencies, percentages, means, standard deviations) and first-order analysis (i.e., chi-square tests, Fisher’s exact test) were performed. Likewise, t-tests or one-way ANOVA tests were performed to determine significant relations of the mean knowledge and attitudes scores with socio-demographic information. Finally, factors that significantly differed in terms of knowledge and attitudes scores, were included into multivariate linear regression analysis with knowledge and attitudes, respectively as the dependent variables. All statistical tests were considered significant at 95% confidence interval with a *p*-value less than .05.

### Ethical consideration

All procedures of the present study were carried out in accordance with the principles for human investigations (i.e., Helsinki Declaration) and also with the ethical guidelines of the Institutional research ethics. Formal ethics approval was granted by the ethical review board of the university research ethics committee [Ref. No: BBEC, JU/ M 2021 COVID-19/2(2)]. Participants in the study were informed about the procedure and purpose of the study and confidentiality of information provided. All participants consented willingly to be a part of the study during the data collection periods. All data were collected anonymously and analyzed using the coding system.

## Results

### General profile of participants

A total of 1658 complete surveys were included in the final analysis. Of them, 56% were males and the participants’ mean age was 23.17 years (SD = 6.05) ranging from 18-65 years. Most were unmarried (86%) and had university/ higher levels of education (83%). The majority were from nuclear families (80%), were in the upper SES group (40%) and came from urban areas (66%). Approximately a quarter (26%) of participants reported that they did not receive all the necessary vaccines in their lifetime (26%).

### Knowledge about the COVID-19 vaccine

The distribution of each knowledge item about the COVID-19 vaccine are presented in Table 2. The mean score of knowledge was 2.83 (SD = 1.48) out of 5, with an overall correct rate of 57%. The mean score of knowledge was significantly higher among participants who reported having university/ higher levels of education, holding nuclear families, in the upper SES group, living in urban areas, and having previous history of receiving all the necessary vaccines (Table 5). Adjusted multiple regression, using only variables that were statistically significant in bivariate analysis, retained having university/ higher levels of education, holding nuclear families, upper SES and having previous history of receiving all the necessary vaccines as factors significantly associated with knowledge (Table 6). Figure 1 shows the participants’ source of knowledge about the COVID-19 vaccine, which is mainly from mass media, social media and the internet.

**Table 5:** Group difference analysis (bivariate) with knowledge and attitudes scores

| Variables | *Knowledge* |  |  |  | *Attitudes* |  |  |  |
| --- | --- | --- | --- | --- | --- | --- | --- | --- |
|  | Mean | (SD) | t/F | p-value | Mean | (SD) | t/F | p-value |
| **Age** |  |  |  |  |  |  |  |  |
| Young | 2.83 | (1.47) | .070 | .792 | 9.36 | (2.40) | .30 | .582 |
| Adult | 2.80 | (1.56) |  |  | 9.26 | (2.35) |  |  |
| **Sex** |  |  |  |  |  |  |  |  |
| Male | 2.88 | (1.49) | 3.111 | .078 | 9.23 | (2.56) | 5.03 | **.025** |
| Female | 2.75 | (1.47) |  |  | 9.49 | (2.15) |  |  |
| **Marital status** |  |  |  |  |  |  |  |  |
| Unmarried | 2.84 | (1.45) | 1.279 | .258 | 9.39 | (2.38) | 2.93 | .087 |
| Married | 2.73 | (1.63) |  |  | 9.10 | (2.45) |  |  |
| **Education** |  |  |  |  |  |  |  |  |
| College/ below | 2.52 | (1.51) | 15.54 | **<.001** | 9.42 | (2.33) | .34 | .559 |
| University/ higher | 2.89 | (1.46) |  |  | 9.33 | (2.40) |  |  |
| **Family type** |  |  |  |  |  |  |  |  |
| Nuclear | 2.88 | (1.47) | 10.29 | **.001** | 9.39 | (2.34) | 1.83 | .177 |
| Joint | 2.60 | (1.48) |  |  | 9.19 | (2.58) |  |  |
| **Monthly family income** |  |  |  |  |  |  |  |  |
| Lower SES | 2.53 | (1.47) | 15.35 | **<.001** | 9.41 | (2.31) | 3.00 | .050 |
| Middle SES | 2.80 | (1.51) |  |  | 9.50 | (2.37) |  |  |
| Upper SES | 3.03 | (1.42) |  |  | 9.18 | (2.46) |  |  |
| **Residence** |  |  |  |  |  |  |  |  |
| Rural | 2.64 | (1.50) | 13.50 | **<.001** | 9.50 | (2.28) | 3.46 | .063 |
| Urban | 2.92 | (1.46) |  |  | 9.27 | (2.44) |  |  |
| **Have you received all the necessary vaccines in your lifetime?** |  |  |  |  |  |  |  |  |
| Yes | 2.94 | (1.45) | 29.14 | **<.001** | 9.43 | (2.34) | 5.64 | **.018** |
| No | 2.50 | (1.51) |  |  | 9.11 | (2.52) |  |  |

**Table 6:** Multivariate regression analysis predicting knowledge and attitudes towards the COVID-19 vaccine

| Variables | *Knowledge<sup>†</sup>* |  |  |  |  | *Attitudes<sup>‡</sup>* |  |  |  |  |
| --- | --- | --- | --- | --- | --- | --- | --- | --- | --- | --- |
| | B | SE | $\beta$ | *t* | *p*-value | B | SE | $\beta$ | *t* | *p*-value |
| Sex<sup>a</sup> | — | — | — | — | — | .272 | .118 | .057 | 2.311 | .021 |
| Education<sup>b</sup> | .34 | .09 | .09 | 3.61 | <.001 | — | — | — | — | — |
| Family type<sup>c</sup> | -.26 | .09 | -.07 | -2.96 | .003 | — | — | — | — | — |
| Monthly family income<sup>d</sup> | .18 | .05 | .10 | 3.87 | <.001 | — | — | — | — | — |
| Residence<sup>e</sup> | .14 | .08 | .05 | 1.79 | .074 | — | — | — | — | — |
| Vaccination history<sup>f</sup> | -.38 | .08 | -.11 | -4.61 | <.001 | -.325 | .133 | -.060 | -2.440 | .015 |

Note: B=unstandardized regression coefficient; SE=Standard error; $\beta$ =standardized regression coefficient;
<sup>a</sup>1=Male, 2=Female; <sup>b</sup>1= College/ below, 2= University/ higher; <sup>c</sup>1= Nuclear, 2= Joint; <sup>d</sup>1= Lower SES, 2= Middle SES, 3= Upper SES; <sup>e</sup>1= Rural, 2= Urban; <sup>f</sup>1=Yes, 2=No;
<sup>†</sup> $F_{(6,1651)} = 14.75$ ; $p < .001$ , $R^2_{Adj} = .047$ ;
<sup>‡</sup> $F_{(12,326)} = 5.50$ ; $p = .004$ , $R^2_{Adj} = .005$ .

**Figure 1:**
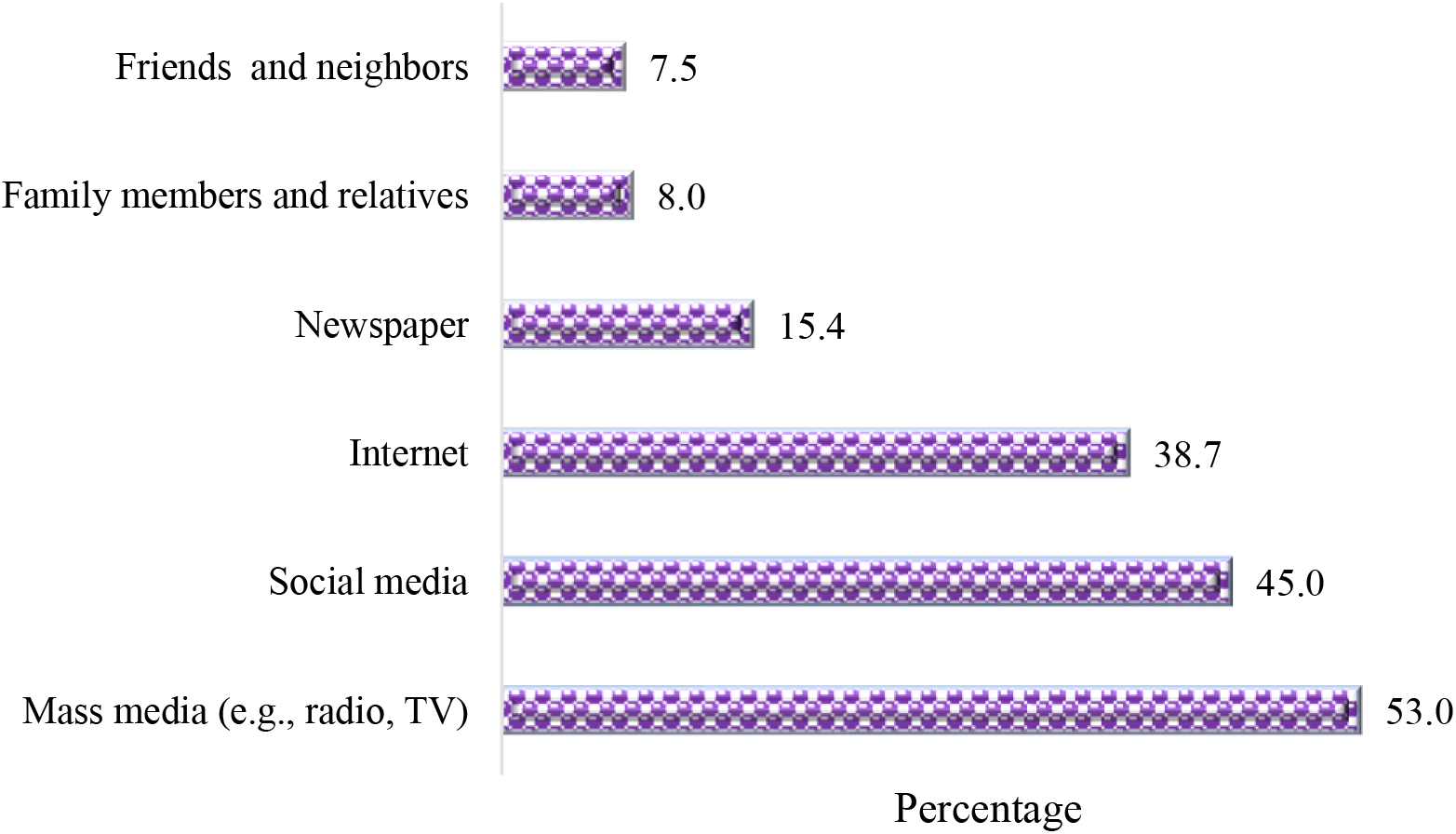
Participants’ source of knowledge about COVID-19 vaccine

### Attitudes towards the COVID-19 vaccine

The distribution of each of the attitudes items towards the COVID-19 vaccine are presented in Table 3. The mean score of attitudes was 9.34 (SD = 2.39) out of 12, with an overall ‘positive attitude’ score of 78%. The mean score of attitudes was significantly higher among participants who reported being female and having previous history of receiving all the necessary vaccines (Table 5). Adjusted multiple regression, using only variables that were statistically significant in bivariate analysis, retained being female and having previous history of receiving all the necessary vaccines as factors significantly associated with attitudes (Table 7). Of particular interest is that only about a quarter of participants (26%) regard the current COVID-19 vaccine in Bangladesh as safe, almost 60% would have the vaccine without hesitation and two-thirds would encourage family or friends to have the vaccination. Therefore, public health work is required to positively impact on attitudes to the COVID-19 vaccination.

### Perceptions towards the COVID-19 vaccine

The distribution of each perceptions item about COVID-19 vaccine are presented in Table 4. With regard to the question ‘*Who should have been vaccinated?*’, just over half (52%) of participants thought everyone should be vaccinated. This perception was significantly higher among females versus males (56.9% vs. 47.9%, *p* = .004). 61% responded that health workers should be vaccinated first, which was significantly higher among females versus males (61.8% vs. 60.2%, *p* = .029). Almost 95% of participants responded that the vaccine should be administered free of charge in Bangladesh, which was significantly higher among females versus males (96.3% vs. 94.1%, *p* = .040). In addition, most participants believed that the newly discovered COVID-19 vaccine may have side effects (89%). Over half of participants (56%) responded that if everyone in the society maintains the preventive measures, the COVID-19 pandemic can be eradicated without vaccination and about a third (35%) responded that they would not purchase the vaccine at their own expense if it was not provided free of charge by the government.

## Discussion

In order to halt the ongoing pandemic, the COVID-19 vaccine has been framed as the ideal solution. Substantial numbers of vaccine candidates are being developed and several clinical trials have recently been released with positive results, leading to a number of countries approving specific vaccines for implementation in vaccination programs. In Bangladesh, the government has already started the COVID-19 vaccination roll-out [23], bringing hope as part of a pandemic solution. Although there are numerous vaccination services in Bangladesh, the complete newness of the COVID-19 vaccination roll-out poses questions about the distribution and acceptance of vaccines in this country. It also poses questions about the general population’s knowledge, attitudes and perceptions towards the COVID-19 vaccine and vaccination rollout. This paper presents findings from a novel study conducted to assess the knowledge, attitudes and perceptions towards COVID-19 vaccinations in Bangladesh. The findings reflect a large number of socio-demographic factors influencing knowledge, attitudes and perceptions towards COVID-19 vaccinations and therefore our findings will be crucial in developing COVID-19 vaccination related awareness and health education programs.

The knowledge regarding COVID-19 vaccinations was low among more than half of the population. In this study, knowledge was significantly associated with education, family type, monthly income of a family, and previous vaccine uptake experience. However, attitudes were significantly associated with only sex and earlier vaccine administration experience. Importantly, the majority of participants (78%) showed positive attitude towards COVID-19 vaccine. Knowledge regarding COVID-19 vaccinations was not significant in terms of participants’ sex. This finding is similar with other studies in Bangladesh demonstrating no significant sex differences in knowledge regarding COVID-19 [24,25]. This finding is similar to studies concerning knowledge towards COVID-19 (not vaccinations) conducted in Bangladesh which reported that males had marginally higher scores in knowledge regarding COVID-19 than females [26,27]. However, this finding is inconsistent to studies concerning knowledge towards COVID-19 (not vaccinations) conducted in Bangladesh which reported that males had marginally higher scores in knowledge regarding COVID-19 than females [26,27]. These discrepancies of knowledge found in our study on COVID-19 vaccinations are possibly due to limited government exposures to information or publicity on COVID-19 vaccinations since the vaccine rollout started. In addition, the potential under-reporting or misinformation of data on the seriousness of incidence and mortality of COVID-19 may reduce concerns about vaccine safety [28] or indeed make residents of Bangladesh reluctant to seek information on either COVID-19 or related vaccinations. Thus, it is essential to support community members by providing easy access to trusted, evidence-based vaccine information.

According to our study, participants with a higher level of education were found to have more knowledge about COVID-19 vaccinations, which is also supported by previous research. Similar scenarios were found in other earlier studies in Bangladesh, illustrating that individuals with a higher educational background showed more knowledge regarding COVID-19 [25,29]. It may be the case that more educated people are more knowledgeable and concerned about their health and well-being, through access to more information sources, and become more engaged in life events that could impact them [30], such as COVID-19 vaccinations.

Significant associations were found between living in a nuclear family and knowledge regarding COVID-19 vaccinations. This is not supported by a preceding study in the same country regarding knowledge about cholera and oral cholera vaccine, where no significant association between knowledge and family type was found [31]. People in the upper SES group had more knowledge regarding COVID-19 vaccinations, which is similar to a study conducted in Indonesia to assess Dengue vaccine acceptance [32]. Moreover, a study carried out among Bangladeshi women regarding knowledge of cervical cancer and HPV vaccine also supports this SES current association [33].

People who have received any vaccine earlier were found to have more knowledge regarding COVID-19 vaccinations in this study. A recent study in China evaluating COVID-19 vaccine acceptance found that people who were previously vaccinated against influenza were more likely to accept the COVID-19 vaccine [34], which was also demonstrated in a study in Hong Kong [7]. This tendency among people may be due to previous positive experiences from vaccination. The level of knowledge about COVID-19 vaccinations were significantly higher among people living in the urban areas, compared to rural areas. However, this association didn’t retain in multiple regression. This is supported by an earlier study in Bangladesh which demonstrated significant correlation between COVID-19 knowledge and urban location [26]. However, our finding is inconsistent with a recent study which found more accurate knowledge about COVID-19 among people in rural areas in Bangladesh [24].

In the present study, 78% of participants had more positive attitudes towards COVID-19 vaccine, which was significantly associated with being female. This association is in line with a previous study on attitudes towards dengue vaccination conducted in Indonesia [35] and attitudes towards COVID-19 carried out in Bangladesh [24]. Attitude towards COVID-19 vaccinations in another study found that female’s hesitation towards COVID-19 vaccinations was higher than that for males [36]. However, a study in China found that male participants were more likely to accept the COVID-19 vaccine [34]. Worldwide, the there is a broadly gendered notion that women’s positive attitudes towards preventive measures are higher than men [24], and this result can be important because engaging women in household-level education and encouragement of COVID-19 vaccination could eventually lead to the improvement in vaccination programs. In addition, In our study, participants who received all the recommended vaccines earlier in life showed more positive attitudes towards COVID-19 vaccinations. Our finding is similar to a study in Hong Kong which demonstrated that the key factors influencing the willingness to receive the A/H7N9 influenza vaccine have been previous vaccine uptake history [7], which is also supported by an earlier study in China [34].

Our study found that over half (52%) of the participants thought that everyone should get COVID-19 vaccine in Bangladesh. Furthermore, more than half of the participants (61%) thought health workers should get the vaccine first. This perception might be because health care workers are at the frontline to diagnose with COVID-19 as they have to deal with patients regularly. In contrast to the general population, the probability of reporting a positive COVID-19 test was higher for frontline health workers [37]. Moreover, their family members are also at high risk, which may lead to rapid transmission among communities.

In our study, 89% of participants assumed that the recently discovered COVID-19 vaccine (the vaccine currently being used in Bangladesh) could have some side-effect, which is similar to a study in the US [36]. A study in China found that 48% of respondents postponed vaccination before confirmation of the safety of the vaccine [34], which shows their doubt regarding vaccine safety. Worryingly, the exceptionally rapid pace of vaccine development, the skepticism of certain groups of science and health experts might elevate doubt about COVID-19 vaccine [38]. According to our study, 95% participants reported that the vaccine should be free for people in Bangladesh, which is inconsistent with a previous study in Indonesia, where most of the respondents were willing to pay for the COVID-19 vaccine [39]. Another study also demonstrated that participants were willing to pay from US$11-US$ for the COVID-19 vaccine [40]. Additionally, a survey in Ecuador also showed 85% of the respondents were willing to pay for COVID-19 vaccination [41]. This difference from the other countries may reflect the financial condition of a large proportion Bangladeshi people, compounded by the fact that lots of people have lost their livelihood during COVID-19 lockdowns and quarantines, which has also contributed to unemployment, hunger, starvation and social disputes [42] which resulted in low ability to pay for COVID-19 vaccine among mass people.

### Limitations

There are certain limitations that should be addressed in the interpretation of the findings from this study. Firstly, the study was cross-sectional, so the causality cannot be attributed to the findings in the regression models. In this respect, a longitudinal study is important. Secondly, the study used an online self-reporting method that may be exposed to social acceptability and memory biases. However, the survey was performed prior to the mass vaccination period in Bangladesh, and its findings may vary after the vaccination program has been implemented. Nevertheless, this is the first study of community perceptions about COVID-19 vaccinations in Bangladesh and will be crucial for health policy makers and planners who are aiming to vaccinate the highest proportion of the population possible in order to mitigate the impacts of the pandemic.

## Conclusions

The COVID-19 pandemic continues to wreak global havoc on lives and livelihoods, and the COVID-19 vaccine represents a possible light of hope for the future. The present study revealed inadequate knowledge but more positive attitudes towards COVID-19 vaccinations in Bangladesh. The findings suggest immediate health education programs and more accurate information should be distributed and advertised by respective health authorities. Policy makers should take steps to ensure adequate knowledge, positive attitudes and perceptions towards COVID-19 vaccinations in order to reduce the vaccine hesitancy facilitated and encouraged by misinformation in the media.

## Data Availability

The datasets used and/or analyzed during the current study are available from the corresponding author on reasonable request.

## Acknowledgments

Firstly, the authors would like to express the most profound gratitude to all of the respondents who participated in this study. The authors would like to thank all research assistants: Mst. Shauda Ahmed, Jannatul Mawa, Amena Akhter, Md. Mehedy Hassan, Sharin Siddika Chaity, Md. Jahid Hasan, Md Tariq Ahmed, Mintia Islam Austha, Md. Mirazul Islam, Md. Al Amin, Asif Mahmud Arnob, Shazid Hossain Adib, Mashrafi Nayeem, Tanoy Chandra Bhowmick, Irfanul Haque Nabil, Mst. Jemi Hkatun, Md. Kamrul Hasan Shaowkat, Sheikh Hafsa Boqul, Shahrina Tasnim Manami, Sangita Chakraborty, Nowshin Nabila Siddique, Imtiaz Ahmed Sakib, and Umma Motahara, for their supports during the study periods.

