## Supplementary file for "Knowledge, attitudes and perceptions towards COVID-19 vaccinations: a cross-sectional community survey in Bangladesh"

| Informed consent | |
|---|---|
| Greetings, dear we have started a survey entitled "Knowledge, attitudes and perceptions towards COVID-19 vaccinations: a cross-sectional community survey in Bangladesh". The results of the study will represent the Knowledge, attitudes and perceptions towards COVID-19 vaccinations. The survey may take 10-15 minutes. Nowhere in the survey, you won't be asked for your personal information. All of your information will be kept secret. You have the right to participate or deny, and during the time of participation, you can withdraw yourself from responding. The study will not be benefitted you by money or other compensations but the outcome of the study may consider by the policymakers and take initiative for COVID-19 vaccinations in Bangladesh. The participants below 18 years should not take part in the survey. | |
| Are you willing to participate in the survey? | o Yes<br>o No |
| **Section 1: Socio-demographic information** | |
| 1. Age | ……………………….. |
| 2. Sex | o Male<br>o Female |
| 3. Marital status | o Unmarried<br>o Married |
| 4. Education | o College/ below<br>o University/ higher |
| 5. Family type | o Nuclear<br>o Joint |
| 6. Monthly family income | o <15,000 Bangladeshi Taka (BDT)<br>o 15,000-30,000 BDT<br>o >30,000 BDT |
| 7. Residence | o Rural<br>o Urban |
| 8. Have you received all the necessary vaccines in your lifetime? | o Yes<br>o No |
| **Section 2: Knowledge** | |
| 1. Do you know about the COVID-19 vaccine? | o Yes<br>o No<br>o Don't know |
| 2. Do you know about the effectiveness of COVID-19 vaccine? | o Yes<br>o No<br>o Don't know |
| 3. Is it dangerous to use overdose vaccines? | o Yes<br>o No<br>o Don't know |
| 4. Does vaccination increase allergic reactions? | o Yes<br>o No |

| | |
|---|---|
| | o Don't know |
| 5. Does vaccination increase autoimmune diseases? | o Yes<br>o No<br>o Don't know |
| 6. How you came to know about COVID-19 vaccines first? | o Mass media (e.g., radio, TV)<br>o Social media (e.g., Facebook, Twitter)<br>o Internet<br>o Newspaper<br>o Family members and relatives<br>o Friends and neighbors |
| **Section 3: Attitudes** | |
| 1. The newly discovered Covid-19 vaccines are safe. | o Disagree<br>o Undecided<br>o Agree |
| 2. The COVID-19 vaccines are essential for us. | o Disagree<br>o Undecided<br>o Agree |
| 3. I will take the COVID-19 vaccine without any hesitation, if it is available in Bangladesh | o Disagree<br>o Undecided<br>o Agree |
| 4. I will also encourage my family/friends/ relatives to get vaccinated. | o Disagree<br>o Undecided<br>o Agree |
| 5. It is not possible to reduce the incidence of COVID-19 without vaccination. | o Disagree<br>o Undecided<br>o Agree |
| 6. The COVID-19 vaccine should be distributed fairly to all of us. | o Disagree<br>o Undecided<br>o Agree |
| **Section 4: Perceptions** | |
| 1. Do you think the newly discovered Covid-19 vaccine may have side effects? | o Yes<br>o No |
| 2. Do you think that if everyone in the society maintains the preventive measures, the COVID-19 pandemic can be eradicated without Vaccination? | o Yes<br>o No |
| 3. Who should have been vaccinated, do you think? | o Those who have not yet been infected with COVID-19<br>o People infected with COVID-19<br>o Newly recovered from COVID-19<br>o Everyone |
| 4. Who's supposed to be vaccinated first, you think? | o General public<br>o Health worker<br>o Public/ private employee<br>o Teacher/ student |

| | |
|---|---|
| | ○ Garment worker<br>○ Businessman |
| 5. Do you think the vaccine should be administered free of charge in Bangladesh? | ○ Yes<br>○ No |
| 6. Would you buy the vaccine at your own expense if it was not provided free by the government? | ○ Yes<br>○ No |
